# Serological Responses to the First Three Doses of SARS-CoV-2 Vaccination in Inflammatory Bowel Disease: A Prospective Cohort Study

**DOI:** 10.1101/2022.03.16.22272440

**Authors:** Joshua Quan, Christopher Ma, Remo Panaccione, Lindsay Hracs, Nastaran Sharifi, Michelle Herauf, Ante Markovinović, Stephanie Coward, Joseph W. Windsor, Léa Caplan, Richard J. M. Ingram, Jamil N. Kanji, Graham Tipples, Jessalyn K. Holodinsky, Charles N. Bernstein, Douglas J. Mahoney, Sasha Bernatsky, Eric I. Benchimol, Gilaad G. Kaplan, the STOP COVID-19 in IBD Research Group

## Abstract

**Background:** Individuals with inflammatory bowel disease (IBD) who are immunocompromised may have a reduced serological response to the SARS-CoV-2 vaccine. We investigated serological responses following 1^st^, 2^nd^, and 3^rd^ doses of SARS-CoV-2 vaccination in those with IBD.

**Methods:** A prospective cohort study of persons with IBD (*n* = 496) assessed serological response 1–8 weeks after 1^st^ dose vaccination, 1–8 weeks after 2^nd^ dose, 8 or more weeks after 2^nd^ dose, and at least 1 week after 3^rd^ dose. Seroconversion and geometric mean titer (GMT) with 95% confidence intervals (CI) were assessed for antibodies to the SARS-CoV-2 spike protein. Multivariable linear regression models assessed the adjusted fold change (FC) in antibody levels.

**Results:** Seroconversion and GMT increased from post-1^st^ dose to 1–8 weeks post-2^nd^ dose (81.6%, 1814 AU/mL vs. 98.7%, 9229 AU/mL, *p*<0.001), decreased after 8 weeks post-2^nd^ dose (94.9%, 3002 AU/mL, *p*<0.001), and rebounded post-3^rd^ dose (99.6%, 14639 AU/mL, *p*<0.001). Prednisone was the only IBD-related medication associated with diminished antibody response after 3^rd^-dose vaccination (FC: 0.07 [95% CI: 0.02, 0.20]). Antibody levels steadily decline following the 2^nd^ (FC: 0.92 [95% CI: 0.90, 0.94] per week) and 3^rd^ dose (FC: 0.88 [95% CI: 0.84, 0.92] per week) of the SARS-CoV-2 vaccine.

**Conclusion:** A three-dose regimen of vaccination to SARS-CoV-2 yields a robust antibody response for those with IBD across all classes of IBD therapies except for prednisone. The decaying antibody levels following the 3^rd^ dose of the vaccine should be monitored in future studies.

## Background

Individuals with inflammatory bowel disease (IBD) may have an increased risk of infection and complications of coronavirus disease 2019 (COVID-19) because of disease activity and/or an immunocompromised state due to their medical therapy. Highly effective vaccines against Severe Acute Respiratory Syndrome Coronavirus 2 (SARS-CoV-2) have been made widely available to reduce disease severity of COVID-19.^1^ The most commonly used vaccines in Canada have been: Pfizer-BioNTech BNT162b2 mRNA (Comirnaty®), NIH-Moderna mRNA-1273 (Spikevax®), and AstraZeneca ChAdOx1 nCoV-19 (Vaxzevria®) adenovirus vector vaccine, which were initially administered as two-dose regimens.^2-4^

Several studies in the IBD population have demonstrated adequate serological response to the two-dose regimen of vaccines.^5^ However, a reduced antibody response has been reported for those on certain treatments, including: tumor necrosis factor antagonists (anti-TNF), immunomodulators, tofacitinib, oral corticosteroids, and combinations of immunosuppressive therapies.^6-11^ Moreover, the decay of antibody levels has been shown in the IBD population, with therapies such as anti-TNF associated with a more rapid decline.^7^ Consequently, gastroenterology organizations and public health officials have recommended three-dose vaccine regimens for individuals with IBD.^12^ Despite recommendations, uptake of 3^rd^ dose vaccines remains low in the IBD population.^13^

We examined the serological response following the 1^st^, 2^nd^, and 3^rd^ dose of SARS-CoV-2 vaccines in those with IBD; the decay of antibodies over time; and the factors, including medications, associated with antibody titres in those with IBD.

## Methods

### Patients and Recruitment

Serological Testing to Outline Protocols for COVID-19 in Inflammatory Bowel Disease (STOP COVID-19 in IBD) is a prospective, observational cohort study of adults with IBD who are followed for serological response to vaccination against SARS-CoV-2. Participants were recruited from October 22, 2020, to September 15, 2021 based on the following inclusion criteria: 1. age 18 years or older, 2. a confirmed diagnosis of IBD, 3. Alberta resident managed by a gastroenterologist in the Calgary Health Zone, and 4. received at least one dose of a SARS-CoV-2 vaccine.

All participants provided informed consent, and the study was approved by the University of Calgary’s Conjoint Health Research Ethics Board (REB20-1082). Reporting was conducted according to STROBE guidelines for reporting observational studies.

### Vaccination and Serology

Serum was processed at the Public Health Laboratory (Alberta Precision Laboratories [APL]) for SARS-CoV-2 serology testing. The Abbott SARS-CoV-2 IgG Architect assay was used to detect antibodies to the nucleocapsid protein of SARS-CoV-2 (anti-N). Nucleocapsid antibody levels were assessed for serosurveillance of natural infection to SARS-CoV-2 with seroconversion defined as an anti-N antibody concentration of ≥0.7 signal/cutoff index.^1^ Anti-N antibody levels were measured to assess a prior infection with SARS-CoV-2.

COVID-19 vaccines also generate an antibody response to the receptor-binding domain (RBD) of the S1 subunit; thus, antibodies to the spike protein can be caused by natural infection or vaccination. Antibodies to the RBD of the S1 subunit of the spike protein (anti-S) were assessed using the Abbott Architect SARS-CoV-2 IgG II Quant assay (Abbott, Abbott Park, Illinois, USA).^14^ The threshold for antibody positivity was defined as ≥50 arbitrary units per millimetre (AU/mL).^15^ The sensitivity and specificity of the SARS-CoV-2 IgG II Quant assay was 98.1% and 99.6%, respectively.^16^ Additionally, antibody levels are reported in the World Health Organization binding antibody units (BAU) to allow for comparison across assays.^17,18^

Serum samples were drawn for assessment of anti-S and anti-N antibodies to SARS-CoV-2 following the 1^st^, 2^nd^, and/or 3^rd^ dose of a SARS-CoV-2 vaccine. Samples were stratified into four groups based on availability of serum samples following vaccination: A. 1–8 weeks following the 1^st^ dose of the vaccine, B. 1–8 weeks following the 2^nd^ dose of the vaccine, C. >8 weeks following the 2^nd^ dose of the vaccine, and D. at least one week following the 3^rd^ dose of the vaccine. Additional samples following 3^rd^ dose vaccination were collected when available but were excluded from the primary analysis; these data were only used for a sub-analysis of within-individual comparisons.

### Outcomes

The primary outcomes of interest were anti-S antibody concentration 1–8 weeks following the 1^st^ dose of a vaccine, 1–8 weeks following the 2^nd^ dose of a SARS-CoV-2 vaccine, >8 weeks following the 2^nd^ dose, and >1 week following the 3^rd^ dose of a vaccine. Secondarily, we assessed the proportion of participants who seroconverted following each dose of vaccine as defined by a threshold ≥50 AU/mL.

### Variables

Study coordinators performed medical chart reviews, and participants completed self-report electronic questionnaires. Age, sex, IBD type (Crohn’s disease versus ulcerative colitis or IBD type unclassified), and IBD medications were collected. Age at date of collection was defined as both a continuous variable and a binary variable stratified as 18–65 years versus >65 years. IBD medications were classified in the following mutually exclusive groups: A. No immunosuppressive therapies including no medical treatment, use of 5-ASA (oral or topical), and/or oral budesonide; B. immunomodulator monotherapy (azathioprine, 6-mercaptopurine, or methotrexate); C. anti-TNF monotherapy (infliximab, adalimumab, or golimumab, originator or biosimilar); D. vedolizumab monotherapy; E. ustekinumab monotherapy; F. tofacitinib monotherapy; G. combination therapy, defined as any combination of two or more of anti-TNF, immunomodulators, vedolizumab, ustekinumab, or tofacitinib; or H. oral corticosteroids, defined as prednisone at any dose alone or in conjunction with any other medication. Medication status was reviewed at each dose of the vaccine to allow for change of therapy through course of vaccination schedules. Medications were classified for each of the four vaccine groups (i.e., post-1^st^ dose, 1–8 weeks post-2^nd^ dose, 8+ weeks post-2^nd^ dose, and post-3^rd^ dose).

Vaccine type and date of vaccination for each dose was recorded. Mixing of vaccine types was not an exclusion criterion. Participants were stratified based on vaccine schedules between 1^st^, 2^nd^, and 3^rd^ doses. For timing between 1^st^ and 2^nd^ doses, “on schedule” was defined as 3–6 weeks between dose administration, and “delayed schedule” was defined as >6 weeks between doses. For timing between 2^nd^ and 3^rd^ doses, “on schedule” was defined as 4–18 weeks between dose administration, and “delayed schedule” was defined as >18 weeks between doses.

Prior history of COVID-19 was defined by a molecular-confirmed diagnosis of SARS-CoV-2 infection via PCR, collected from chart review. Among individuals who did not have a confirmed PCR test, we also defined prior exposure to SARS-CoV-2 as those who had an anti-N seroconversion.

Finally, for each of the four vaccine groups (i.e., post-1^st^ dose, 1–8 weeks post-2^nd^ dose, 8+ weeks post-2^nd^ dose, and post-3^rd^ dose), we established the time from vaccine dose in weeks.

### Statistics

Statistical analyses were performed using STATA v.16.0 (StataCorp) with a significance threshold of 0.05 and two-sided testing. Geometric mean titres (GMT) with associated 95% confidence intervals (CI) or standard deviation (SD) were used to report anti-S concentrations. Seroconversion rates were reported as a proportion of individuals with positive seroconversion. Mann-Whitney U tests were performed to compare GMTs between vaccine dose categories. Additional sensitivity analyses were performed with Wilcoxon signed rank tests to compare GMTs between vaccine dose categories using paired data only.

Univariable associations of anti-S concentrations were determined using two-tailed, unpaired t-tests comparing log-transformed antibody titres within demographic, disease, and vaccine-related variables for each vaccine dose category. Multivariable linear regression models were used for each vaccine dose group (i.e., post-1^st^ dose, 1–8 weeks post-2^nd^ dose, 8+ weeks post-2^nd^ dose, and post-3^rd^ dose) to determine the effect of independent predictors on log-transformed anti-S concentration. Sex, age at time of collection, disease type, medication class, prior COVID-19 history, delayed vaccination schedule between doses, and time after vaccination were identified *a priori* as independent predictors in our primary models. Exponentiated coefficients represented the fold change (FC) associated with each binary covariate and the FC in log anti-S concentration per unit change for continuous variables.^7^

Within-individual comparisons of antibody titres were assessed into subset populations: A. Individuals with serum samples in both the post 1–8 weeks and 8+ weeks 2^nd^ dose groups; B. serum samples following 1–8 weeks after 2^nd^ dose and 3^rd^ dose vaccine; C. serum samples following 8+ weeks after 2^nd^ dose and 3^rd^ dose vaccine; and D. individuals with multiple serum samples following the 3^rd^ dose of the vaccine. Mean within-individual differences in antibody concentration were calculated for each subset and stratified by prior infection to SARS-CoV-2. For each subset, overall and stratified mean antibody titres were compared between time-points using paired t-tests.

We conducted a sensitivity analysis whereby age was modelled as a binary variable (18– 65 years versus >65 years) instead of a continuous variable. We also conducted a subgroup analysis of individuals who received either the Pfizer-BioNTech or Moderna vaccines without mixing of an mRNA vaccine. In this subgroup, we included a binary variable comparing the Pfizer-BioNTech and Moderna vaccines within the multivariable regression model. In all secondary and sensitivity analyses, models were adjusted for the same covariates defined in the primary model.

### Data Visualization

The 3D plots were created in R (R Core Team) using the rgl: 3D Visualization Using OpenGL package. Data points were plotted with days from vaccine dose on the x-axis, the patient’s age at time of collection on the y-axis, and antibody levels on the z-axis (converted to a logarithmic scale). The regression plane in each plot depicts a linear regression model carried out with antibody level as the outcome and days from vaccine dose and age at time of collection as predictors.^19^

Our data are available to view in an open access, online, interactive dashboard (https://kaplan-gi.shinyapps.io/COVID_Serology/) created in R (R Core Team) using the Shiny: Web Application Framework for R^3^ package.^20^

## Results

Table 1 describes the distribution of demographic, disease, and vaccine-related characteristics for the entire cohort (*n* = 496) and for individuals with serological data 1–8 weeks following 1^st^ dose (*n* = 250), 1–8 weeks following 2^nd^ dose (*n* = 315), 8+ weeks following 2^nd^ dose (*n* = 237) and 1+ weeks following 3^rd^ dose vaccination (*n* = 232).

**Table 1.** Patient characteristics per vaccine dose category

| Characteristic | Overall | Post-1<sup>st</sup> Dose<br>(1–8 weeks) | Post-2<sup>nd</sup> dose<br>(1–8 weeks) | Post-2<sup>nd</sup> dose<br>(8+ weeks) | Post-3<sup>rd</sup> dose<br>(1+ weeks) |
| --- | --- | --- | --- | --- | --- |
| **Patients, *n*** | 496 | 250 | 315 | 237 | 232 |
| **Sex, *n* (%)** |  |  |  |  |  |
| Male | 226 (45.6) | 117 (46.8) | 146 (46.4) | 99 (41.8) | 99 (42.7) |
| Female | 270 (54.4) | 133 (53.2) | 169 (53.6) | 138 (58.2) | 133 (57.3) |
| **Mean age (SD)** | 49.7 (14.7)<sup>a</sup> | 50.2 (14.7) | 50.1 (14.6) | 52.5 (14.8) | 52.7 (14.0) |
| **IBD Type, *n* (%)** |  |  |  |  |  |
| Crohn's Disease | 360 (72.6) | 192 (76.8) | 220 (69.8) | 171 (72.1) | 180 (77.6) |
| Ulcerative Colitis | 125 (25.2) | 52 (20.8) | 87 (27.6) | 63 (26.6) | 50 (21.5) |
| IBD-U | 11 (2.22) | 6 (2.4) | 8 (2.6) | 3 (1.3) | 2 (0.9) |
| **Medication, *n* (%)** |  |  |  |  |  |
| No immunosuppressives | 44 (8.9) | 29 (11.6) | 38 (12.1) | 26 (11.0) | 16 (6.9) |
| Anti-TNF only | 173 (34.9) | 70 (28.0) | 106 (33.7) | 85 (35.8) | 90 (38.8) |
| Immunomodulators only | 11 (2.2) | 5 (2.0) | 6 (1.9) | 3 (1.3) | 6 (2.6) |
| Vedolizumab only | 50 (12.1) | 31 (12.4) | 44 (14.0) | 34 (14.3) | 21 (9.0) |
| Ustekinumab only | 108 (21.8) | 60 (24.0) | 64 (20.3) | 54 (22.8) | 45 (19.4) |
| Tofacitinib only | 6 (1.2) | 3 (1.2) | 5 (1.6) | 3 (1.3) | — |
| Combination therapy* | 79 (15.9) | 46 (18.4) | 45 (14.3) | 27 (11.4) | 48 (20.7) |
| Oral Corticosteroids** | 15 (3.0) | 6 (2.4) | 7 (2.2) | 5 (2.1) | 6 (2.6) |
| **Vaccine Type, *n* (%)** | — |  |  |  |  |
| Pfizer | — | 200 (80.0) | 257 (81.6) | 203 (85.7) | 211 (91.0) |
| Moderna | — | 40 (16.0) | 51 (16.1) | 29 (12.2) | 21 (9.0) |
| AstraZeneca | — | 10 (4.0) | 7 (2.2) | 5 (2.1) | — |
| **Seroconversion, *n/N* (%)** | — | 204/250 (81.6) | 311/315 (98.7) | 225/237 (94.9) | 231/232 (99.6) |
| No Prior COVID-19 | — | 117/223 (79.4) | 284/288 (98.6) | 193/205 (94.2) | 209/210 (99.5) |
| Prior COVID-19 | — | 27/27 (100.0) | 27/27 (100.0) | 32/32 (100.0) | 22/22 (100.0) |
| **GMT (95% CI)** | — | 1814 (981, 2648) | 9229 (7871, 10588) | 3002 (2222, 3783) | 14639 (12846, 16472) |
| No Prior COVID-19 | — | 494 (373, 616) | 8414 (7137, 9690) | 2020 (1539, 2501) | 13736 (11898, 15573) |
| Prior COVID-19 | — | 12715 (6058, 19372) | 17931 (10121, 25740) | 9293 (4794, 13792) | 23470 (16372, 30569) |

<sup>a</sup> For overall sample, age represents age at 1<sup>st</sup> dose vaccination
\*Combination therapy refers to any combination of two or more of the following therapies: anti-TNF, immunomodulators, vedolizumab, ustekinumab, and tofacitinib.
\*\*Oral prednisone at any dose or with any other drug class.

Seroconversion rates were 81.6% (1^st^ dose, 1–8 weeks), 98.7% (2^nd^ dose, 1–8 weeks), 94.9% (2^nd^ dose, 8+ weeks), and 99.6% (3^rd^ dose, 1+ weeks) with 100% seroconversion for those with a prior COVID-19 infection for all dose categories (Table 1).

GMT levels significantly increased (*p*<0.001) from 1–8 weeks after 1^st^ dose (1814 AU/mL [95% CI: 981, 2648 AU/mL]) to 1–8 weeks after 2^nd^ dose (9229 AU/mL [95% CI: 7871, 10588 AU/mL]), but fell significantly (*p*<0.001) to 3002 AU/mL (95% CI: 2222, 3783 AU/mL) 8+ weeks after 2^nd^ dose vaccination (Table 1, Figure 1). Following 3^rd^ dose vaccination, antibody levels increased significantly compared to all vaccine categories (*p*<0.001) to 14639 AU/mL (95% CI: 12846, 16472 AU/mL). The distribution of individual antibody concentrations per vaccine dose category is displayed in Figure 1.

**Figure 1.**
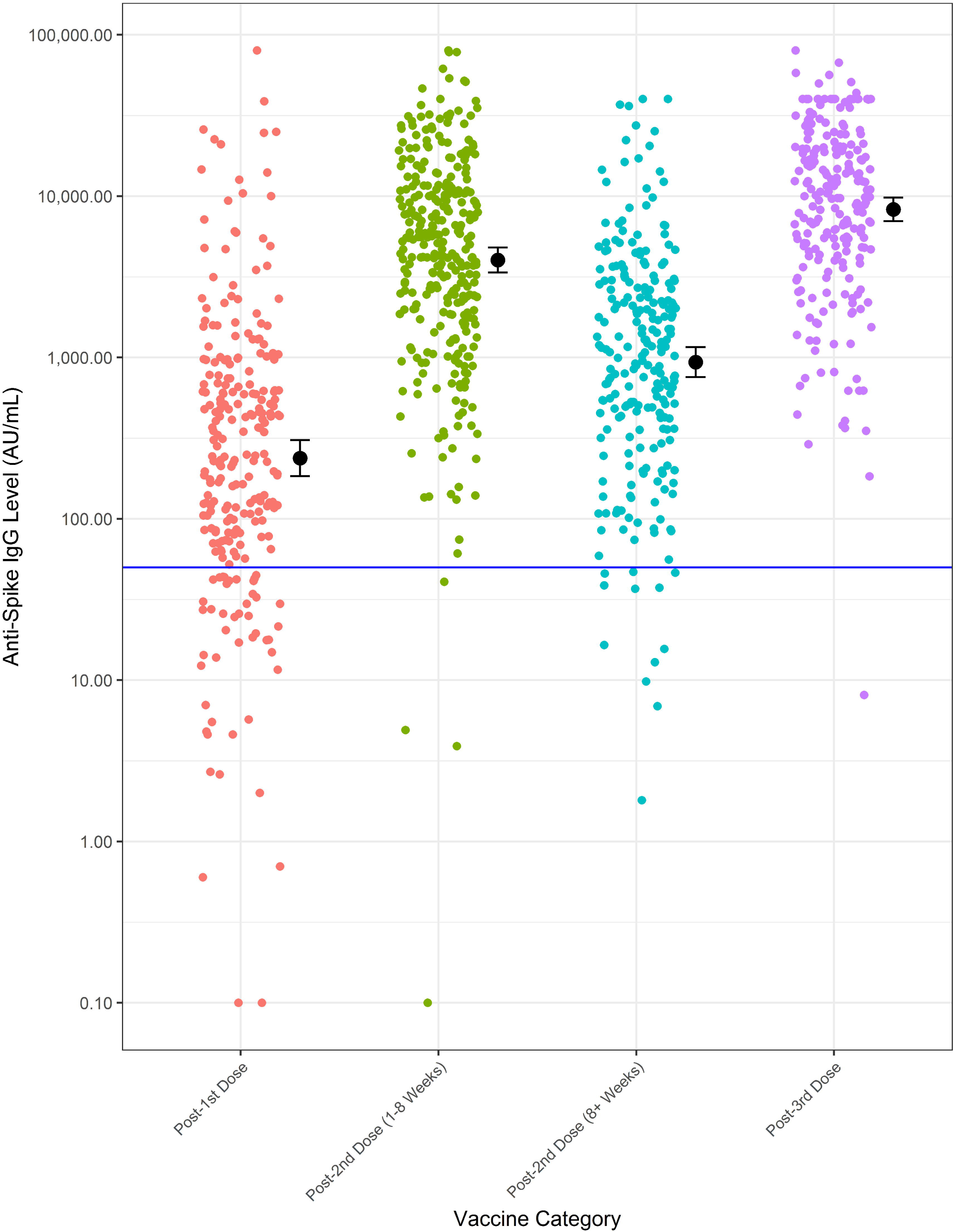
Anti-SARS-CoV-2 spike antibody concentration per vaccine category. Black points represent GMTs while narrow black bars represent bounds of 95% CI associated with each GMT. Solid blue line represents threshold for positive seroconversion (50 AU/mL). Serological data from this study are available open access on an online interactive ShinyApp: https://kaplan-gi.shinyapps.io/COVID_Serology/

Age at time of serology was independently associated with decreased log-transformed anti-S concentration for all vaccine dose groups except post-3^rd^ dose. FC values indicate factor of log anti-S concentration per decade increase in age, for example log anti-S concentration decreased by 0.71 (95% CI: 0.64, 0.80) per decade increase in age for samples taken 8+ weeks post-2^nd^ dose vaccine. Three-dimensional plots displaying anti-S concentration by age and days after vaccination indicate decreasing titres with increasing age and time after vaccination for 2^nd^ dose, whereas for 3^rd^ dose, titres only decrease with time after vaccination (Video 1 and Video 2). Age >65 years was associated with lower GMT for all vaccine dose groups except post-3^rd^ dose (Supplemental Table 1).

For each vaccine dose category, patients on corticosteroids had the lowest GMT compared to other medication classes (Table 2, Supplemental Table 2). Multivariable linear regression analyses demonstrated a significant association between corticosteroid use and decreased log-transformed anti-S concentration compared to individuals not on immunosuppressive medication, across all vaccine dose categories (1–8 weeks post-2^nd^ dose FC: 0.07 (95% CI: 0.02, 0.20). Anti-TNF monotherapy and combination therapy were also predictors of decreased anti-S titres for all vaccine dose categories except post-3^rd^ dose vaccination (Table 2). The FC in log anti-S concentration compared to the no immunosuppressive medication group for each medication is displayed in Table 2. For example, at 1–8 weeks post-2^nd^ dose, anti-TNF monotherapy was associated with a decrease in log anti-S concentration of 0.56 (95% CI: 0.33, 0.94) compared to individuals without immunosuppressive medications.

**Table 2.** Stratified geometric mean titres and exponentiated coefficients of multivariate linear regression model per each vaccine dose category.

| Variable | Post-1<sup>st</sup> dose<br>(n = 250) |  | Post-2<sup>nd</sup> Dose (1–8weeks)<br>(n = 315) |  | Post-2<sup>nd</sup> dose (8+ weeks)<br>(n = 237) |  | Post- 3<sup>rd</sup> dose<br>(n = 232) |  |
| --- | --- | --- | --- | --- | --- | --- | --- | --- |
|  | GMT | Fold change (95% CI) | GMT | Fold change (95% CI) | GMT | Fold change (95% CI) | GMT | Fold change (95% CI) |
| **Age<sup>a</sup>**<br>Per decade | — | **0.85 (0.80, 0.89)** | — | **0.78 (0.70, 0.87)** | — | **0.71 (0.64, 0.80)** | — | 0.91 (0.82, 1.01) |
| **Sex**<br>Male<br>Female | 1524<br>2070 | 0.78 (0.52, 1.17) | 8163<br>10151 | 0.85 (0.63, 1.16) | 3115<br>2922 | 0.87 (0.63, 1.19) | 12947<br>15933 | 0.97 (0.72, 1.31) |
| **IBD Type**<br>Crohn's Disease<br>Ulcerative Colitis/IBD-U | 1698<br>2198 | 0.71 (0.42, 1.20) | 8693<br>10379 | 0.79 (0.55, 1.13) | 2639<br>3945 | 0.82 (0.57, 1.19) | 14835<br>14049 | 0.87 (0.62, 1.23) |
| **Medication**<br>No immunosuppressives* | 2138 | — | 11514 | — | 3049 | — | 13556 | — |
| Anti-TNF only | 2743 | 0.69 (0.33, 1.45) | 6460 | **0.56 (0.33, 0.94)** | 2127 | **0.22 (0.13, 0.37)** | 12861 | 0.72 (0.40, 1.29) |
| Immunomodulators only | 210 | 0.44 (0.09, 2.07) | 4149 | 0.44 (0.14, 1.44) | 13543 | 0.26 (0.06, 1.09) | 13904 | 0.76 (0.27, 2.12) |
| Vedolizumab only | 1755 | 1.56 (0.68, 3.57) | 15152 | 1.53 (0.84, 2.80) | 4171 | 1.69 (0.92, 3.12) | 18326 | 1.52 (0.74, 3.11) |
| Ustekinumab only | 1568 | 1.45 (0.68, 3.10) | 13532 | 1.54 (0.87, 2.72) | 3094 | 0.90 (0.51, 1.61) | 21038 | 1.61 (0.86, 3.01) |
| Tofacitinib only | 8427 | 0.97 (0.14, 6.80) | 9596 | 0.53 (0.15, 1.88) | 5598 | 0.40 (0.09, 1.73) | — | — |
| Combination therapy | 517 | **0.19 (0.08, 0.42)** | 3555 | **0.22 (0.12, 0.40)** | 2978 | **0.18 (0.09, 0.35)** | 12449 | 0.61 (0.32, 1.15) |
| Corticosteroids | 149 | **0.22 (0.05, 0.92)** | 2756 | **0.07 (0.02, 0.20)** | 946 | **0.03 (0.01, 0.09)** | 2320 | **0.07 (0.02, 0.20)** |
| **Vaccine Schedule**<br>Scheduled<br>Delayed | N/A<br>N/A | — | 9309<br>9192 | 0.75 (0.54, 1.04) | 2264<br>3621 | **0.52 (0.38, 0.72)** | 13420<br>15922 | 0.81 (0.61, 1.09) |
| **Prior COVID-19**<br>Yes<br>No | 12714<br>494 | **32.56 (16.95, 62.55)** | 17931<br>8414 | **2.44 (1.43, 4.15)** | 9293<br>2020 | **4.65 (2.92, 7.38)** | 23470<br>13736 | **1.97 (1.22, 3.20)** |
| **Weeks After Vaccine<sup>b</sup>** | — | 0.93 (0.80, 1.07) | — | **0.82 (0.75, 0.89)** | — | **0.92 (0.90, 0.94)** | — | **0.88 (0.84, 0.92)** |

\* Indicates reference group
<sup>a</sup> Values represent change in anti-S concentration per decade increase in age
<sup>b</sup> Values represent change in anti-S concentration per week increase in duration after vaccination

Prior COVID-19 history was significantly associated with increased log anti-S concentration across all vaccine dose categories with the largest FC associated with post 1^st^ dose responses (FC=21.91 [95% CI: 16.37, 54.64]) and the lowest impact associated with post 3^rd^ dose responses (FC=1.97 [95% CI: 1.22, 3.20]). Delayed 2^nd^ dose vaccination schedule (>6 weeks between 1^st^ and 2^nd^ doses) was associated with increased log anti-S concentration 8+ weeks following 2^nd^ dose vaccination (*p*<0.001). However, the effect of delayed 3^rd^ dose vaccination schedule (>18 weeks between 2^nd^ and 3^rd^ doses) was not observed.

Antibody titres decreased significantly (*p*<0.001) following 2^nd^ dose vaccination; Figure 2a demonstrates an inverse trend between anti-S antibody concentrations and days after 2^nd^ dose vaccination. Figure 2b. presents a similar trend for post-3^rd^ dose results, with overall lower anti-S antibody concentrations for serology samples taken further from the 3^rd^ dose vaccination date.

**Figure 2a.**
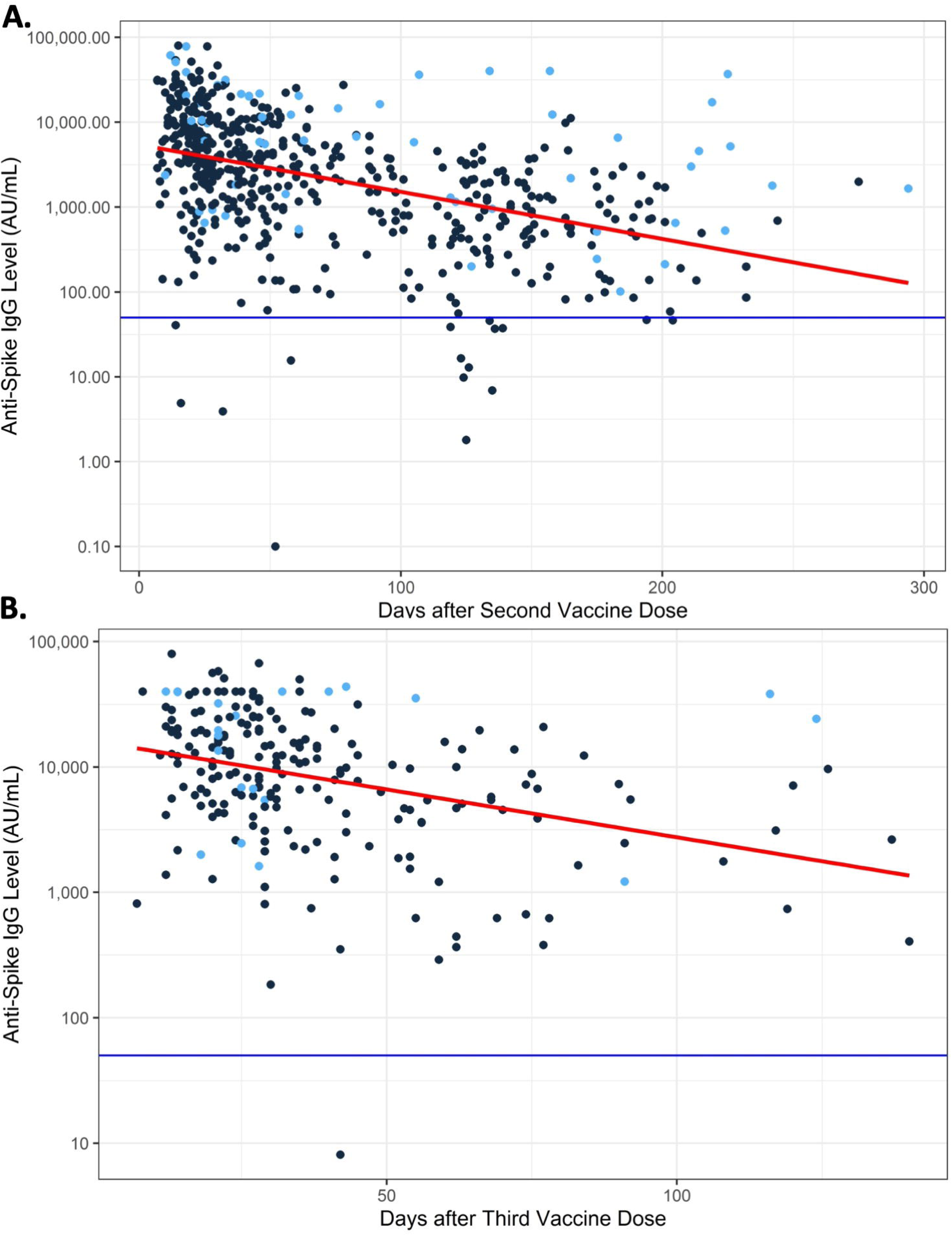
Anti-S concentration per days after dose vaccination. Figure 2b. Anti-S concentration per days after 3^rd^ dose vaccination. Blue points represent individuals with prior SARS-CoV-2 infection. Solid red lines represent line of best fit for linear relationship between anti-S concentration (AU/mL) and time after vaccination. Solid blue lines represents threshold for positive seroconversion (50 AU/mL). Serological data from this study are available open access on an online interactive ShinyApp: https://kaplan-gi.shinyapps.io/COVID_Serology/

Multivariable regression analyses identified a significant relationship between weeks after vaccination and log anti-S concentration for 1–8 weeks post-2^nd^ dose (FC: 0.82 [95% CI: 0.75, 0.89] per week), 8+ weeks post-2^nd^ dose (FC: 0.92 [95% CI: 0.90, 0.94] per week), and 1+ weeks post-3^rd^ dose (FC: 0.88 [95% CI: 0.84, 0.92] per week) groups. Antibodies decayed by a factor of 0.88 or 12% per week (95% CI: 8% to 16%) following the 3^rd^ dose of vaccine.

The mean difference in antibody concentration from post 1–8 weeks to 8+ weeks after 2^nd^ dose (*n*=124) was -7668 AU/mL (95% CI: -10055, -5281 AU/mL, *p*<0.001). After stratifying by prior SARS-CoV-2 infection, differences were only significant for individuals without prior infection (−6970 AU/mL [95% CI: -8999, -4941 AU/mL]). The individual rate of decay in individuals without prior SARS-CoV-2 infection was determined to be 396 AU/mL per week (95% CI: 277, 516 AU/mL per week).

GMT following post-3^rd^ dose vaccination increased with a mean difference in antibody titres between post-3^rd^ dose and 1–8 weeks post-2^nd^ dose vaccination (*n* = 153) of 8148 AU/mL (95% CI: 6155, 10141 AU/mL, *p*<0.001). Within-individual comparison following 8+ weeks post-2^nd^ dose and post-3^rd^ dose vaccination (*n* = 99) also demonstrated that GMT following post-3^rd^ dose vaccination increased significantly with a mean difference of 12400 AU/mL (95% CI: 9281, 15518 AU/mL) (Table 3).

**Table 3.** Mean individual differences from within individual comparisons for each population subset.

| Characteristic | Post 2<sup>nd</sup> (1–8 Weeks) vs.<br>Post- 2<sup>nd</sup> (8+ Weeks) | Post-2<sup>nd</sup> (1–8 Weeks) vs.<br>Post-3<sup>rd</sup> Dose | Post-2<sup>nd</sup> (8+ Weeks) vs.<br>Post-3<sup>rd</sup> Dose | Within Post-3<sup>rd</sup> dose |
| --- | --- | --- | --- | --- |
| **Number of patients, *n*** | 124 | 153 | 99 | 28 |
| Prior COVID-19 | 10 | 12 | 8 | 5 |
| No Prior COVID-19 | 114 | 141 | 91 | 23 |
| **Mean individual difference,**<br>***AU/mL* (95% *CI*)** | –7668 (–10055, –5281) | 8148 (5166, 10141) | 12400 (9281, 15518) | –8057 (–13811, –2303) |
| Prior COVID-19 | –13158 (–28468, 2153) | 12492 (765, 24220) | 18261 (2078, 34443) | –15264 (–55085, 24557) |
| No Prior COVID-19 | –6970 (–8999, –4941) | 7778 (5805, 9752) | 11918 (8699, 15137) | –6490 (–10080, –2901) |

Within-individual comparison of multiple 3^rd^ dose vaccination serology values (*n =* 28) demonstrated a mean decrease in individual anti-S concentration of 8057 AU/mL (95% CI: 2303, 13811 AU/mL) between earlier and later collection dates. The median duration between serology collection dates was 13.5 weeks (IQR: 11.1–15.7 weeks). The individual rate of decay following 3^rd^ dose vaccination in individuals without prior infection to SARS-CoV-2 was determined to be 483 AU/mL per week (95% CI: 237, 729 AU/mL per week).

Sensitivity analyses comparing paired data between vaccine dose categories did not indicate any differences in statistical significance. Sensitivity analyses modelling age as a binary variable (18–65 years versus >65 years) also demonstrated significant associations with increased age (>65 years) and decreased antibody response for post-1^st^ dose, 1–8 weeks post-2^nd^ dose, and 8+ weeks post-2^nd^ dose. Likewise, there was no association for post-3^rd^ dose responses (*p*=0.579) (Supplemental Table 1). Subgroup analyses excluding non-mRNA vaccination and vaccine mixing did not impact the significance of covariate associations with log anti-S concentration. Subgroup models for post 1^st^ dose and 1–8 weeks post-2^nd^ dose responses indicate significantly decreased log anti-S concentration for Pfizer compared to Moderna (FC: 0.41 [95% CI: 0.24, 0.70]; FC: 0.51 [95% CI: 0.34, 0.78]) (Supplemental Table 3). Supplemental Table 4 reports antibody levels in the World Health Organization BAU/ml.

## Discussion

A three-dose SARS-CoV-2 vaccination schedule for individuals with IBD has been proposed based on impaired serological responses to two-dose regimens.^12,21,22^ Our study demonstrates near complete seroconversion to 3^rd^ dose vaccination with significantly higher antibody titres when compared to 2^nd^ dose vaccination. Third dose antibody responses are consistently high across all IBD therapies for maintenance of remission including biologic and immunomodulator therapies. In contrast, those with IBD taking prednisone at the time of receiving a vaccine mounted lower antibody responses following all three doses. Despite robust initial antibody responses to vaccination, our study indicates decay of antibody titres over time following vaccination for both 2^nd^ and 3^rd^ doses. While these data support widespread implementation of a three-dose vaccine regimen, future studies are necessary to determine if additional doses are required to maintain sufficient antibody levels over time.

Our findings indicate a limited antibody response to a single dose of a SARS-CoV-2 vaccine, but consecutively stronger responses following each additional dose. Only 81.6% of individuals with IBD seroconverted following the 1^st^ dose of a SARS-CoV-2 vaccine. However, 98.7% of participants seroconverted following two doses with a corresponding five-fold increase in mean antibody titres. Similarly, a meta-analysis in the IBD population showed an increase from 49.2% to 93.7% seroconversion after 1^st^ and 2^nd^ doses, respectively.^5^ Our study demonstrated a substantial increase in serological response to a 3^rd^ dose of a vaccine, which was similar to an American cohort of IBD patients that demonstrated a robust antibody response following an extra dose from the initial vaccine series.^23^ Consistent seroconversion and antibody titres responses across different geographic jurisdictions and using different antibody assays highlights the replicative external validity of the strong antibody response to three dose regimens of the SARS-CoV-2 vaccines in the IBD population.

While considerably large serological responses were observed immediately after 2^nd^ and 3^rd^ doses, the linear regression models showed that antibody titres diminished over time after vaccination. Within-individual analyses also indicated that antibody levels decayed at an approximate rate of 400 AU/mL per week after 2^nd^ dose and 500 AU/mL per week after 3^rd^ dose. Decay of antibody titres following vaccine doses has been previously reported;^7,24^ however, 3^rd^ dose antibody titres diminish from a considerably higher baseline as compared to the 2^nd^ dose vaccine. The threshold level for antibody titres to offer protection from an infection to SARS-CoV-2 is not known. A prior study in the UK demonstrated that following the 2^nd^ dose of the vaccine that a 10-fold increase in antibody titres was associated with 0.8-fold decrease in breakthrough infections.^7^ Future studies are necessary to establish the antibody level necessary to confer protection to SARS-CoV-2 following three vaccine doses.

Older age was associated with decreased antibody titres after 1^st^ and 2^nd^ doses in prior studies,^6-8^ which is clinically relevant as seniors with IBD are at highest susceptibility to severe COVID-19.^25^ A prior study showed that antibody titres fell by 21% per decade of life following the 2^nd^ dose of vaccine.^6^ Similarly, our models also indicated decreases in antibody concentration per decade of life across 1^st^ and 2^nd^ doses. For example, immediately after 2^nd^ dose of the vaccine, antibody titres were 12% lower per decade of advancing age (Video 1). However, our study presents novel data on the effect of age on 3^rd^ dose responses and indicates no change in antibody titres per decade of advancing age (FC: 0.91 [95% CI: 0.82, 1.01]). These data highlight the fact that seniors with IBD mount lower antibody responses after a two-dose regimen but mount comparably high antibody responses following a three-dose regime; thus, the importance of a 3^rd^ dose for sufficient immune protection in seniors with IBD.

We assessed infection to SARS-CoV-2 prior to each dose of vaccine by verifying either a PCR-confirmed molecular diagnosis and/or positive serosurveillance with nucleocapsid antibodies. Our data were consistent with prior studies showing that COVID-19 infection before vaccination increases antibody levels following each dose of the vaccine.^7,8^ Whereas prior infection induced a 33-fold increase in 1^st^ dose antibody responses, exposure to SARS-CoV-2 diminished to only a 2.0-fold increase for 3^rd^ dose responses. We used a lower threshold for a positive anti-N level (i.e., 0.7) than the manufacturer recommended level of 1.4, which is supported by serosurveillance studies of SARS-CoV-2.^26^

Our data are consistent with previous studies in demonstrating diminished antibody responses following two doses of a SARS-CoV-2 vaccine for patients on anti-TNF therapy, combination immunosuppressive therapies, and corticosteroids.^6-11,24,27,28^ In contrast, our study showed that the 3^rd^ dose of the vaccine yielded high antibody titres across all classes of medications used for maintenance of remission in IBD. However, administration of vaccine while concurrently taking prednisone yielded lower antibody response following 1^st^, 2^nd^, and 3^rd^ dose of the vaccine. While prednisone was associated with diminished responses, mean antibody titres following 3^rd^ dose vaccination were still 2.5 times higher than those more than eight weeks following 2^nd^ dose in patients on oral corticosteroids. Findings across medication class should be interpreted cautiously where small sample size reduces the precision of estimates. For example, small sample size limited our ability to analyze the effect of prednisone across different dosages (e.g., above and below 20 mg). While our data highlight that all individuals with IBD should receive a 3^rd^ dose of the vaccine, regardless of medication class, those on prednisone at the time of receiving a vaccine may ultimately require four doses.

Several limitations of our study should be considered. Due to a low number of patients with breakthrough COVID-19 infections, we were unable to assess the effectiveness of SARS-CoV-2 vaccination or evaluate whether antibody titres were associated with breakthrough infections. In addition, we did not assess other immune responses to SARS-CoV-2 vaccination, including neutralizing antibodies or T-cell immunity, which may serve as more functional proxies of immune protection over antibody titres.^24,29^ Due to logistical constraints in data collection during the pandemic, we were unable to obtain samples corresponding to all vaccine doses for all patients, resulting in semi-independent vaccine dose groups. While a community-based recruitment strategy involving all gastroenterologists in the Calgary zone was utilized, population-based sampling was not possible. Further, we did not compare those with IBD to healthy controls; however, a recent serology study performed in the general population within the same jurisdiction and using the same antibody assay demonstrated higher antibodies than our IBD population after 1^st^ and 2^nd^ dose of the vaccine.^30^ We used the Abbott Architect SARS-CoV-2 IgG II Quant assay, but also converted antibody levels to the World Health Organization BAU/ml (Supplementary Table 4) to support standardization across assays.^17,18^

Timing and type of vaccination was variable due to changing vaccine availability in Canada. We conducted a sensitivity analysis comparing scheduled versus delayed vaccine timing. Delayed vaccine timing was associated with increased antibody responses following 2^nd^ dose vaccination; however, timing of vaccine schedule did not influence antibody levels after the 3^rd^ dose. Also, most participants received mRNA vaccines, with only 4.0% and 2.2% of patients receiving ChAdOx1 nCoV-19 for 1^st^ and 2^nd^ dose vaccination, respectively. Several participants also received mixed doses of mRNA and ChAdOx1 nCoV-19 or mixed doses of differing mRNA vaccines. However, our sensitivity analysis restricting the sample to those with the same mRNA vaccine across all doses confirmed the consistency of the interpretation of the data.

Our study demonstrated novel findings surrounding antibody responses to 3^rd^ dose vaccination, which indicate the importance toward maintaining humoral immunity across all medication classes and age groups. The one exception was individuals with IBD who were vaccinated while on prednisone, who may require a 4^th^ dose of vaccine in the future. Considering the significant decay of antibodies following a two-dose regimen and robust response following the 3^rd^ dose, healthcare providers and public health officials should prioritize communication of the necessity of a three-dose vaccine regimen to ensure sufficient serological protection to patients with IBD. Finally, future studies should evaluate the decay of antibodies following the 3^rd^ dose to establish whether a fourth dose is ultimately necessary for most individuals with IBD.

## Data Availability

All data produced are available online at https://kaplan-gi.shinyapps.io/COVID_Serology/

## Abbreviations

anti-S: antibodies to the receptor-binding domain of the S1 subunit of the spike protein
anti-N: antibodies to the nucleocapsid protein of SARS-CoV-2
anti-TNF: tumour necrosis factor antagonist
APL: Alberta Precision Laboratories
AU: arbitrary units
CI: confidence interval
COVID-19: coronavirus disease 2019
FC: fold change
GMT: geometric mean titer
IBD: inflammatory bowel disease
RBD: receptor-binding domain
SARS-COV-2: severe acute respiratory syndrome coronavirus 2
SD: standard deviation

