## Supplement for "Serological Responses to the First Three Doses of SARS-CoV-2 Vaccination in Inflammatory Bowel Disease: A Prospective Cohort Study"

**Supplement A.**

*Supplemental Table 1. Stratified geometric mean titres and exponentiated coefficients of multivariate linear regression model per each vaccine dose category with categorical age.*

| **Variable** | **Post-1^st^ dose**  (*n* = 250) | | **Post-2^nd^ Dose (1–8weeks)**  (*n* = 315) | | **Post-2^nd^ dose (8+ weeks)** (*n* = 237) | | **Post-3^rd^ dose**  (*n* = 232) | |
| --- | --- | --- | --- | --- | --- | --- | --- | --- |
|  | GMT | FC (95% CI) | GMT | FC (95% CI) | GMT | FC (95% CI) | GMT | FC (95% CI) |
| **Sex**  Male  Female | 1524  2070 | 0.76 (0.50, 1.14) | 8163  10151 | 0.81 (0.59, 1.11) | 3115  2922 | 0.87 (0.62, 1.21) | 12947  15933 | 0.96 (0.71. 1.29) |
| **Age**  >65 Years  =< 65 Years | 2488  1655 | **0.59 (0.36, 0.99)** | 7462  9628 | **0.61 (0.41, 0.92)** | 1769  3358 | **0.49 (0.33, 0.73)** | 15492  14129 | 0.90 (0.63, 1.29) |
| **IBD Type**  Crohn’s Disease  Ulcerative Colitis/IBD-U | 1698  2198 | 0.68 (0.40, 1.15) | 8693  10379 | 0.73 (0.51, 1.05) | 2639  3945 | 0.85 (0.58, 1.25) | 14835  14049 | 0.85 (0.60, 1.20) |
| **Medication**  No immunosuppressives*  Anti-TNF only  Immunomodulators only  Vedolizumab only  Ustekinumab only  Tofacitinib only  Combination therapy  Corticosteroids | 2138  2743  210  1755  1568  8427  517  149 | —  0.78 (0.37, 1.63)  0.44 (0.09, 2.09)  1.74 (0.76, 4.01)  1.54 (0.72, 3.30)  1.15 (0.16, 8.10)  **0.22 (0.10, 0.48)**  **0.21 (0.05, 0.87)** | 11514  6460  4149  15152  13532  9596  3555  2756 | —  0.65 (0.38, 1.10)  0.51 (0.15, 1.71)  1.76 (0.96, 3.24)  1.35 (0.95, 3.06)  0.59 (0.16, 2.17)  **0.27 (0.14, 0.50)**  **0.07 (0.02, 0.22)** | 3049  2127  13543  4171  3094  5598  2978  946 | —  **0.24 (0.14, 0.43)**  0.38 (0.08, 1.72)  1.83 (0.96, 3.48)  1.36 (0.51, 1.71)  0.55 (0.12, 2.52)  **0.22 (0.11, 0.44)**  **0.03 (0.01, 0.09)** | 13556  12861  13904  18326  21038  —  12449  2320 | —  0.75 (0.41, 1.35)  0.80 (0.29, 2.27)  1.51 (0.73, 3.11)  1.64 (0.87, 3.08)  —  0.66 (0.35, 1.25) **0.07 (0.02, 0.20)** |
| **Vaccine Schedule**  Scheduled  Delayed | N/A  N/A | — | 9309  9192 | 0.76 (0.55, 1.07) | 2264  3621 | **0.52 (0.37, 0.72)** | 13420  15922 | 0.84 (0.63, 1.13) |
| **Prior COVID-19**  Yes  No | 12714  494 | **31.78 (16.47, 61.31)** | 17931  8414 | **2.39 (1.39, 4.12)** | 9293  2020 | **5.21 (3.21, 8.47)** | 23470  13736 | **1.93 (1.19, 3.14)** |
| **Weeks After Vaccine**  Per week | — | 0.93 (0.80, 1.08) | — | **0.82 (0.75, 0.89)** | — | **0.88 (0.60, 1.29)** | — | **0.88 (0.85, 0.92)** |

*Supplemental Table 2. Seroconversion rate and GMTs with 95% Cis per medication class*

| **Medication Class** | **Post-1^st^ dose** | | **Post-2^nd^ Dose (1–8weeks)** | | **Post-2^nd^ dose (8+ weeks)** | | **Post-3^rd^ dose** | |
| --- | --- | --- | --- | --- | --- | --- | --- | --- |
|  | SC Rate n/N (%) | GMT (SE) | SC Rate n/N (%) | GMT (SE) | SC Rate n/N (%) | GMT (SE) | SC Rate  n/N (%) | GMT (SE) |
| No immunosuppressives | 26/29 (89.7) | 2138 (1107) | 38/38  (100.0) | 11514 (2043) | 26/26  (100.0) | 3049 (960) | 16/16  (100.0) | 13556 (2540) |
| Anti-TNF only | 59/70  (84.3) | 2743 (1303) | 105/106  (99.1) | 6460 (872) | 80/85  (94.1) | 2127 (666) | 6/6  (100.0) | 13904 (5710) |
| Immunomodulators only | 4/5  (80.0) | 210 (124) | 6/6  (100.0) | 4149 (1720) | 3/3  (100.0) | 13543 (13229) | 90/90  (100.0) | 12861 (1483) |
| Vedolizumab only | 23/31  (90.3) | 1755 (808) | 44/44  (100.0) | 15153 (2344) | 34/34  (100.0) | 4171 (842) | 21/21  (100.0) | 18326 (3615) |
| Ustekinumab only | 54/60 (90.0) | 1568 (406) | 64/64  (100.0) | 13532 (1853) | 54/54  (100.0) | 3094 (668) | 45/45  (100.0) | 21038 (2065) |
| Tofacitinib only | 3/3  (100.0) | 8427 (8129) | 5/5  (100.0) | 9596 (6186) | 3/3  (100.0) | 5598 (4499) | **–** | **–** |
| Combination therapy | 27/46  (58.7) | 517 (159) | 44/45  (97.8) | 3556 (1005) | 23/27  (85.2) | 2978 (1344) | 48/48 (100.0) | 12449 (1846) |
| Corticosteroids | 3/6  (50.0) | 149 (106) | 5/7  (71.4) | 2737 (1171) | 2/5  (40.0) | 946 (877) | 5/6  (83.3) | 2320 (1326) |

*Supplemental Table 3. Stratified geometric mean titres and exponentiated coefficients of multiple linear regression model per each vaccine dose category for individuals with constant mRNA vaccine type.*

| **Variable** | **Post-1^st^ dose**  (*n* = 240) | | **Post-2^nd^ Dose (1–8weeks)**  (*n* = 294) | | **Post-2^nd^ dose (8+ weeks)** (*n* = 224) | | **Post-3^rd^ dose**  (*n* = 178) | |
| --- | --- | --- | --- | --- | --- | --- | --- | --- |
|  | GMT | FC (95% CI) | GMT | FC (95% CI) | GMT | FC (95% CI) | GMT | FC (95% CI) |
| **Age ^a^**  Per decade | — | **0.83 (0.72, 0.96)** | — | **0.82 (0.74, 0.90)** | — | **0.72 (0.64, 0.80)** | — | 0.95 (0.86, 1.05) |
| **Sex**  Male  Female | 1588  2142 | 0.79 (0.53, 1.20) | 7888  10416 | 0.89 (0.65, 1.20) | 3140  3014 | 0.87 (0.62, 1.20) | 12848  16104 | 0.95 (0.71, 1.28) |
| **IBD Type**  Crohn’s Disease  Ulcerative Colitis/IBD-U | 1774  2234 | 0.69 (0.41, 1.17) | 8946  10053 | 0.79 (0.55, 1.12) | 2749  3857 | 0.92 (0.63, 1.33) | 14762  14456 | 0.79 (0.56, 1.11) |
| **Medication**  No immunosuppressives*  Anti-TNF only  Immunomodulators only  Vedolizumab only  Ustekinumab only  Tofacitinib only  Combination therapy  Corticosteroids | 2267  2905  210  1812  1618  8427  527  149 | —  0.83 (0.39, 1.79)  0.47 (0.10, 2.18)  1.92 (0.83, 4.46)  1.69 (0.78, 3.66)  1.20 (0.18, 8.17)  **0.19 (0.09, 0.44)**  **0.23 (0.06, 0.96)** | 12443  6598  4149  14272  14113  9596  3581  3216 | —  **0.54 (0.32, 0.91)**  0.36 (0.12, 1.13)  1.53 (0.83, 2.79)  1.53 (0.86, 2.73)  0.41 (0.12, 1.40)  **0.21 (0.11, 0.39)**  **0.13 (0.04, 0.41)** | 3232  2229  13543  3926  3232  5598  2982  946 | —  **0.21 (0.12, 0.38)**  0.26 (0.06, 1.13)  1.63 (0.86, 3.08)  0.87 (0.47, 1.60)  0.40 (0.09, 1.79)  **0.17 (0.08, 0.34)**  **0.03 (0.01, 0.09)** | 13061  13060  13904  16763  21056  —  12881  3432 | —  0.83 (0.45, 1.53)  0.82 (0.30, 2.24)  1.57 (0.74, 3.31)  1.72 (0.90, 3.32)  —  0.72 (0.37, 1.38)  0.27 (0.08, 0.88) |
| **Vaccine Type**  Pfizer  Moderna | 2002  1294 | **0.41 (0.24, 0.70)** | 8353  14553 | **0.51 (0.34, 0.78)** | 3156  2409 | 0.87 (0.53, 1.43) | 14653  15108 | 0.60 (0.35, 1.03) |
| **Vaccine Schedule**  Scheduled  Delayed | —  — | — | 8614  9620 | **0.68 (0.49, 0.94)** | 2170  3885 | **0.52 (0.37, 0.73)** | 13624  15849 | 0.82 (0.61, 1.10) |
| **Prior COVID-19**  Yes  No | 12715  511 | **33.90 (17.77, 64.66)** | 15614  8667 | **2.20 (1.31, 3.69)** | 9198  2081 | **4.54 (2.80, 7.37)** | 23470  13644 | **1.92 (1.22, 3.05)** |
| **Weeks After Vaccine ^b^**  Per week | — | 0.88 (0.75, 1.02) | — | **0.84 (0.77, 0.91)** | — | 0.92 (0.90, 0.94) | — | 0.88 (0.85, 0.92) |

* Indicates reference group

**^a^** Values represent change in anti-S concentration per decade increase in age

**^b^** Values represent change in anti-S concentration per week increase in duration after vaccination

*Supplemental Table 4. Overall and stratified geometric mean titres in BAU/mL with 95% CIs for each vaccine dose category*

| **GMT in *BAU/mL***  **(95% CI)** | **Post-1^st^ Dose (1–8 weeks)** | **Post-2^nd^ dose (1–8 weeks)** | **Post-2^nd^ dose (8+ weeks)** | **Post-3^rd^ dose (1+ weeks)** |
| --- | --- | --- | --- | --- |
| Overall | 258 (139, 376) | 1311 (1118, 1503) | 426 (316, 537) | 2079 (1824, 2339) |
| Prior COVID-19 | 70 (53, 87) | 1195 (1013, 1376) | 287 (219, 355) | 1951 (1690, 2211) |
| No Prior COVID-19 | 1806 (860, 2751) | 2526 (1437, 3655) | 1320 (681, 1958) | 3333 (2325, 4341) |

**Supplement B.** STROBE Statement—checklist of items that should be included in reports of observational studies

|  | Item No | Recommendation | Page |
| --- | --- | --- | --- |
| **Title and abstract** | 1 | (*a*) Indicate the study’s design with a commonly used term in the title or the abstract | 1 |
|  |  | (*b*) Provide in the abstract an informative and balanced summary of what was done and what was found | 4 |
| Introduction | | |  |
| Background/rationale | 2 | Explain the scientific background and rationale for the investigation being reported | 5 |
| Objectives | 3 | State specific objectives, including any prespecified hypotheses | 5 |
| Methods | | |  |
| Study design | 4 | Present key elements of study design early in the paper | 6 |
| Setting | 5 | Describe the setting, locations, and relevant dates, including periods of recruitment, exposure, follow-up, and data collection | 6 |
| Participants | 6 | (*a*) *Cohort study*—Give the eligibility criteria, and the sources and methods of selection of participants. Describe methods of follow-up  *Case-control study*—Give the eligibility criteria, and the sources and methods of case ascertainment and control selection. Give the rationale for the choice of cases and controls  *Cross-sectional study*—Give the eligibility criteria, and the sources and methods of selection of participants | 6 |
|  |  | (*b*) *Cohort study*—For matched studies, give matching criteria and number of exposed and unexposed  *Case-control study*—For matched studies, give matching criteria and the number of controls per case | N/A |
| Variables | 7 | Clearly define all outcomes, exposures, predictors, potential confounders, and effect modifiers. Give diagnostic criteria, if applicable | 7-8 |
| Data sources/ measurement | 8* | For each variable of interest, give sources of data and details of methods of assessment (measurement). Describe comparability of assessment methods if there is more than one group | 7-8 |
| Bias | 9 | Describe any efforts to address potential sources of bias | 10 |
| Study size | 10 | Explain how the study size was arrived at | 6 |
| Quantitative variables | 11 | Explain how quantitative variables were handled in the analyses. If applicable, describe which groupings were chosen and why | 9 |
| Statistical methods | 12 | (*a*) Describe all statistical methods, including those used to control for confounding | 9 |
|  |  | (*b*) Describe any methods used to examine subgroups and interactions | 9 |
|  |  | (*c*) Explain how missing data were addressed | 17 |
|  |  | (*d*) *Cohort study*—If applicable, explain how loss to follow-up was addressed  *Case-control study*—If applicable, explain how matching of cases and controls was addressed  *Cross-sectional study*—If applicable, describe analytical methods taking account of sampling strategy | 9, 17 |
|  |  | (*e*) Describe any sensitivity analyses | 10 |

Continued on next page

| Results | | | Page |
| --- | --- | --- | --- |
| Participants | 13* | (a) Report numbers of individuals at each stage of study—eg numbers potentially eligible, examined for eligibility, confirmed eligible, included in the study, completing follow-up, and analysed | 19 |
|  |  | (b) Give reasons for non-participation at each stage | 17 |
|  |  | (c) Consider use of a flow diagram | NR |
| Descriptive data | 14* | (a) Give characteristics of study participants (eg demographic, clinical, social) and information on exposures and potential confounders | 19 |
|  |  | (b) Indicate number of participants with missing data for each variable of interest | 19 |
|  |  | (c) *Cohort study*—Summarise follow-up time (eg, average and total amount) | 19, Fig 2. |
| Outcome data | 15* | *Cohort study*—Report numbers of outcome events or summary measures over time | 11, 19 |
|  |  | *Case-control study—*Report numbers in each exposure category, or summary measures of exposure | N/A |
|  |  | *Cross-sectional study—*Report numbers of outcome events or summary measures | N/A |
| Main results | 16 | (*a*) Give unadjusted estimates and, if applicable, confounder-adjusted estimates and their precision (eg, 95% confidence interval). Make clear which confounders were adjusted for and why they were included | 19, 20 |
|  |  | (*b*) Report category boundaries when continuous variables were categorized | 11-21 |
|  |  | (*c*) If relevant, consider translating estimates of relative risk into absolute risk for a meaningful time period | NR |
| Other analyses | 17 | Report other analyses done—eg analyses of subgroups and interactions, and sensitivity analyses | 13-14 |
| Discussion | | |  |
| Key results | 18 | Summarise key results with reference to study objectives | 14 |
| Limitations | 19 | Discuss limitations of the study, taking into account sources of potential bias or imprecision. Discuss both direction and magnitude of any potential bias | 17 |
| Interpretation | 20 | Give a cautious overall interpretation of results considering objectives, limitations, multiplicity of analyses, results from similar studies, and other relevant evidence | 14-18 |
| Generalisability | 21 | Discuss the generalisability (external validity) of the study results | 18 |
| Other information | | |  |
| Funding | 22 | Give the source of funding and the role of the funders for the present study and, if applicable, for the original study on which the present article is based | 2-3 |

*Give information separately for cases and controls in case-control studies and, if applicable, for exposed and unexposed groups in cohort and cross-sectional studies.

**Note:** An Explanation and Elaboration article discusses each checklist item and gives methodological background and published examples of transparent reporting. The STROBE checklist is best used in conjunction with this article (freely available on the Web sites of PLoS Medicine at http://www.plosmedicine.org/, Annals of Internal Medicine at http://www.annals.org/, and Epidemiology at http://www.epidem.com/). Information on the STROBE Initiative is available at www.strobe-statement.org.
