## Supplementary figures and images for "Serological Responses to the First Three Doses of SARS-CoV-2 Vaccination in Inflammatory Bowel Disease: A Prospective Cohort Study"

### Video 1

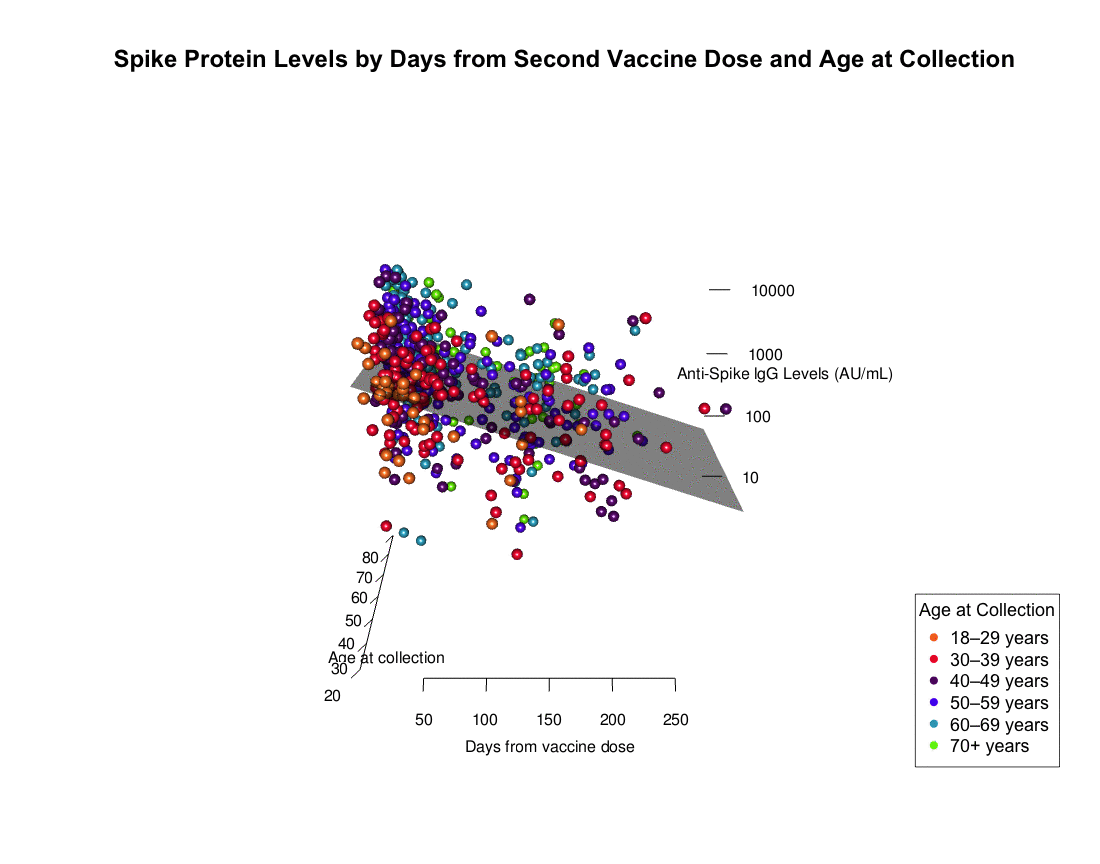

### Video 2

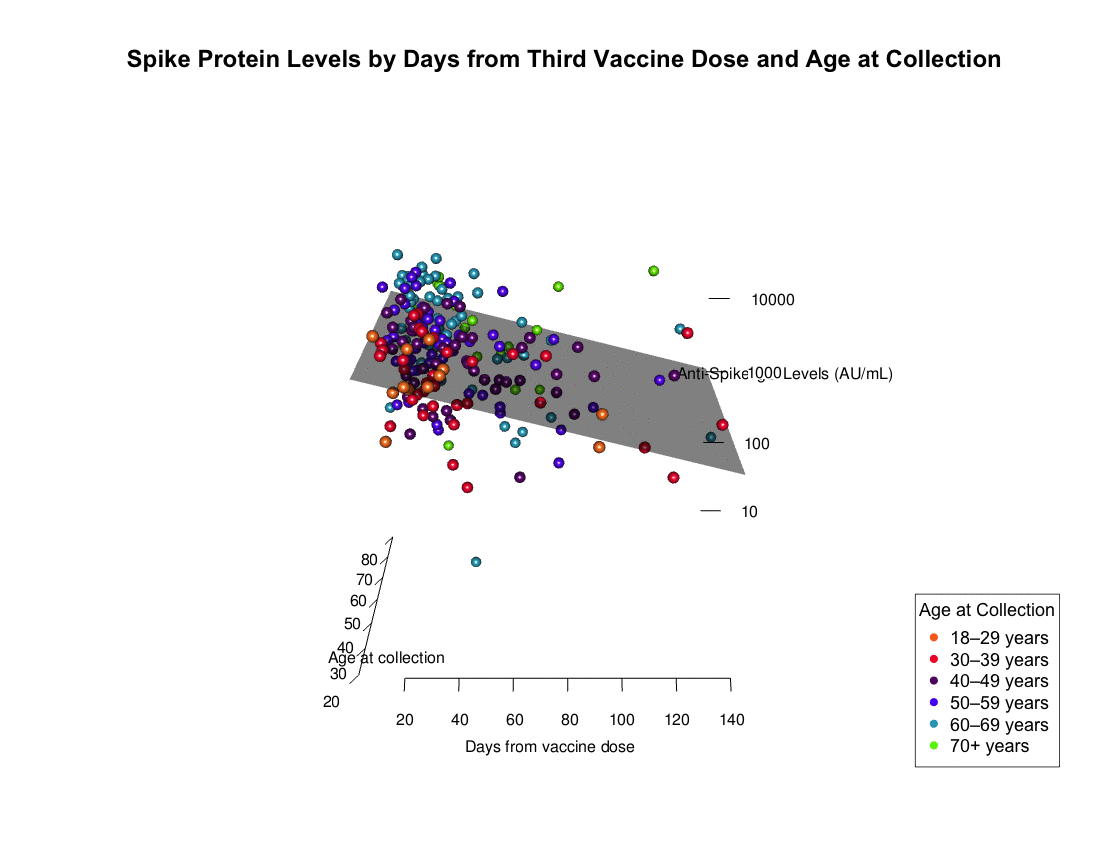
